# Interstitial fluid rejuvenation through young-donor plasma exchange in cognitively impaired patients: a pilot safety and feasibility study

**DOI:** 10.64898/2026.04.29.26351809

**Authors:** Arne Søraas, Andreas Engvig, Dag Alnæs, Lana Nagmadin Karim, Elena Danilova, Mette Stausland Istre, Sara Nygaard, Tone Reinton Utgård, Sandra Ceprnjic, Olaug Reiakvam, Elisabeth Edvardsen, Lars T. Westlye, Anders B. Nygaard, John Arne Dahl, Lise Sofie Haug Nissen-Meyer, Håkon Ihle-Hansen, Petter Holland

## Abstract

Heterochronic parabiosis improves physiological and cognitive function in aging rodents; these benefits appear to be derived from the removal of aged blood plasma components and the addition of younger ones. In humans, removing plasma from older individuals with Alzheimer’s disease (AD) and replacing it with saline and albumin delayed cognitive deterioration in a large clinical trial. However, mimicking heterochronic parabiosis in humans by removing large volumes of a patient’s blood plasma and replacing it with plasma from young and healthy donors has not been tested. Here, we have performed a pilot study to characterize the feasibility and safety of such a procedure, replacing between 16 and 26 L of patient blood plasma with young (ages 18-24) donor blood plasma for twelve patients who recently received a diagnosis of mild cognitive impairment with biomarker evidence of AD. The dose and time interval between plasma exchanges was tailored to maximize equilibration of donor plasma components into the interstitial fluid, achieving what we term interstitial rejuvenation. We explored three permutations of a plasma exchange protocol with different treatment intensities and doses, each performed on three to five patients. We present data on safety, feasibility, patient burden, resource use of the treatments, preliminary measurements of clinical variables and short-term cognitive trajectories in the patients. The procedures were safe and feasible, supporting further investigation of treatment efficacy in a larger controlled trial. This safety and feasibility study was first registered 22 December 2023 at ClinicalTrials.gov and given the identifier NCT06234436.

## Introduction

With heterochronic parabiosis, the circulatory system of an older and a younger animal is directly connected, effectively creating a mixture of young and old blood in both animals. Aging-related physiological benefits of the intervention in the older animals are broad, whereas detrimental effects have been described for the younger animals [1, 2]. While many organs in the older animal show improved physiology due to heterochronic parabiosis, the improvements in cognitive function are most striking. Both beneficial components from younger blood plasma [3, 4] and damaging components in older blood plasma [2, 5] have been found to mediate the cognitive effects by mechanistic studies from several research groups.

We have recently shown that epigenetic aging processes in immune cells accelerate when transplanted into an older body and decelerate in a younger body [6], suggesting that aging processes are modulated by molecular components in the cellular environment, also in humans. An intervention more closely resembling heterochronic parabiosis in humans has not yet been tested. The AMBAR clinical trial did show that removal of older patient blood plasma and replacement by saline and albumin delayed progression of cognitive decline in patients with mild to moderate Alzheimer’s disease (AD), although the effect size was modest [7]. Adding components from young human blood plasma to cognitively impaired patients has been tested, but only with small amounts of blood plasma or extracted blood plasma fractions [8, 9].

AD is a compelling target for young-donor plasma exchange treatment in humans, given the limited availability of disease-modifying treatments. The AMBAR study gave critical information about both benefit from diluting the patient blood plasma with saline and albumin (benefit from removal of “old” components) and demonstrated the safety and feasibility of large volumes of blood plasma exchange in a cognitively impaired patient group. Our hypothesis, based on rodent heterochronic blood exchange studies [10], is that there is additional benefit from adding “young” blood plasma, supporting a treatment modality where patient plasma removal and younger plasma addition are combined. These effects together improve cognitive function in rodents [3], and thus may represent a novel treatment modality to delay cognitive decline in individuals living with MCI.

Here, we describe a human treatment protocol designed to mimic heterochronic parabiosis more closely than prior studies. Our approach entails the exchange of large volumes of an older patient’s plasma with plasma from young and healthy donors. The goal is interstitial fluid rejuvenation, which we define as achieving a molecular composition within organs of the body that more closely resembles that of a 18–24-year-old person compared to the patient’s endogenous molecular environment. The plasma exchange protocol we use is a variation of the standard therapeutic plasma exchange (TPE) procedure, for which safety is well characterized, but mainly in the setting of severe autoimmune diseases. We here tested the safety and feasibility of three different permutations of a treatment protocol, each with different treatment durations and intensity.

## Materials and methods

### Participant selection and recruitment

The study was an open, unblinded study where the patients received standard treatment for early AD led by a memory clinic and/or their general practitioner (GPs). Standard treatment includes diet and exercise advice, addressing risk factors like blood pressure or nutrient deficiencies, and regularly prescribed acetylcholinesterase inhibitors.

Patients were recruited from the outpatient memory clinic at Bærum Hospital, Vestre Viken Hospital Trust, Norway, between 2023 and 2025. Patients referred to memory clinics for suspected AD in Norway are typically younger, often of working age, or present with more complex symptoms, while older individuals are usually managed by their general practitioners (GPs) in primary care. Referrals to memory clinics are made by GPs based on subjective or objective cognitive complaints requiring further evaluation.

Patients were eligible if they had mild cognitive impairment consistent with AD clinically, as well as biomarker evidence of AD pathology, defined as either a positive beta-amyloid (^18^F-flutemetamol) positron emission tomography-computed tomography (PET-CT) scan and/or cerebrospinal fluid (CSF) biomarkers (reduced amyloid-beta 42 and increased total and/or phosphorylated tau levels). Eligible patients had to be in the clinical stage corresponding to MCI, defined according to Winblad criteria [11] as impaired cognitive function with preserved activities of daily living (ADL). There was no upper age limit.

All patients provided written informed consent prior to enrollment. Capacity to consent was evaluated as part of the cognitive assessment before study invitation, and only patients deemed capable were included. The consent form was also signed by the closest relative to all patients. Exclusion criteria were limited life expectancy, a cancer diagnosis within the past 10 years, unwillingness or inability to provide informed consent, or a Montreal Cognitive Assessment (MoCA) score <17.

Plasma donors aged 18-24 were recruited among the regular blood donors and through campaigns in social and regular media. All donors satisfied national requirements for blood donation.

### Safety Committee and study group

A Safety Committee consisting of two senior physicians from the Medical Department at Oslo University Hospital, a cardiologist and an acute medicine specialist were engaged and discussed all adverse events with the principal investigator (PI) and assisted the PI on decisions related to inclusion and continuation of treatments after adverse events.

The clinical study was led by a specialist in internal medicine and infectious diseases together with a study group consisting of a geriatrician and two physician specialists in transfusion medicine.

### Donor blood plasma, donor selection, procedure and plasma testing

According to Norwegian and European regulations, plasma for transfusion is either approved after pathogen reduction or quarantine. In this project, the Directorate of Medicinal Products granted special permission to transfuse single donor, fresh frozen plasma without pathogen reduction or quarantine to project patients, as had also been done in the Norplasma project for Covid-19 convalescent plasma [12]. This is justified by the advantageous epidemiological profile of Norwegian blood donors [13] and the otherwise limited treatment options for the patient group. Patients were informed about these risks as part of the inclusion process described above.

In accordance with blood center Standard Operating Procedures (SOPs), new donors undergo a registration interview with sampling for standard serological testing and blood typing and screening, before approval. New and established donors volunteering to this study were informed about the project and gave written consent to donating plasma and samples for research purposes and use of their deidentified personal information (REK # 520697). 4-6 weeks after registration, 610 ml plasma was harvested with Aurora® plasmapheresis machines (using kits Plasmacell 6R2278, Fresenius Kabi) following blood center SOPs, with sodium citrate 4% (ratio 6) as anticoagulant. For male donors, screening tests for antibodies to human leukocyte antigens (HLA) class I and II were routinely performed at first plasmapheresis procedure. For female donors, anti-HLA screening and human neutrophil antigen 3 typing (HNA3) typing was performed and results obtained before the first plasmapheresis. HNA 3bb-donors were further tested for anti-HNA 3a. Plasma donors having anti-HLA or anti-HNA 3a were excluded from the project when these results were available. In a few cases, units with anti-HLA antibodies were transfused before these results were obtained. Approved plasma donors were invited to repeat donations every 2 weeks. For every 10 donations, we controlled their levels of total protein and IgG, and if these levels dropped below 60 g/L and 6.0 g/L respectively, the donor was referred until protein levels were normalized. Any donor who donated every 2 weeks consecutively for 20 weeks, was paused for at least 1 month. Donors did not receive any payment for their plasma donation beyond a small souvenir given to all blood donors in Norway. At the end of the study, donors received a gift card valued at 50 USD as a thank-you for their participation in the research project.

### Blood plasma exchange procedures, general information

Included patients received plasma exchange in addition to treatment as usual for AD. The plasma exchange procedures were conducted using fresh frozen plasma (FFP) from healthy young (18-24 years old) blood/plasma donors and all FFP was collected at the Blood Bank at Oslo University Hospital. In one instance, a patient received freshly collected plasma (non-frozen). All procedures were conducted in a treatment room equipped to handle anaphylactic reactions and other adverse events (i.e. adrenaline, methylprednisolone, antihistamines, fluid, oxygen, calcium tablets). A defibrillator was stationed outside the treatment room. One to two treatments were conducted simultaneously with one nurse per patient. In addition, a responsible physician was present in the room during treatments and had the responsibility to assist should any acute situation occur and to approve each treatment procedure based on a clinical examination of the patient and recent biochemistry.

The project developed its own treatment SOP (Supplementary X) that was based on the Oslo University Hospital TPE SOP. Key considerations before each treatment were whether the patient was completely free of infection, had not experienced unexpected adverse events since the last treatment and that the most recent set of biochemistry results was normal. Biochemical investigation was conducted approximately every 3-6 procedures.

Every patient procedure was initiated with administration of 500 mg of calcium chewing tablets. Additional calcium chewing tablets were also given before each bag of FFP or if suspected hypocalcaemic reactions occurred. If the patient had experienced a suspected allergic reaction at any previous procedure, 10 mg of the antihistamine cetirizine was also given. Peripheral venous access was secured in both cubital veins, but in some instances other veins, such as on the legs, were used. An 18G intravenous cannula was typically used for both the inlet and return lines; 16G was used for patients with large veins and 20G for the return line when venous access was difficult.

If an allergic reaction occurred, 10 mg cetirizine was given and the physician considered if the procedure should be aborted. During intensive treatment periods, the patients were advised to add extra proteins (eggs or meat) to their diet to compensate for potential loss of proteins during the procedure or from sampling. Patients on ACE inhibitors were not allowed to take their tablets before treatment. Patients were offered a hepatitis B vaccination before treatment.

The patients and their household members were informed about the potentially severe and late-occurring adverse event transfusion-associated lung injury (TRALI) and received a letter with written information about these adverse events. Study staff were always available by phone for 24 hours after each treatment. The letter also contained information the patients could give to healthcare personnel should TRALI occur. For the first treatment session, patients were surveilled for 2 hours after the start of the last plasma transfusion. For other treatment sessions, patients were surveilled for 45 minutes after the start of the last transfusion.

### Blood plasma exchange procedures, “pilot-pilot”

For the first four patients included in the treatment protocol, treatment started with a single bag of FFP and with longer intervals of time between treatments. This was to ensure safety at the lowest doses while also establishing the practicalities related to the treatment protocols. After the safety of one FFP bag was established, some patients received single higher dose treatments of two or three bags of FFP as part of their “pilot-pilot” phase.

### Blood plasma exchange procedures, intensive

An intensive phase was conducted with nine to ten treatments, each with three bags of FFP (each 610 ml) every 2-3 days over a period of approximately four weeks. The procedure was conducted on the Terumo Spectra Optia machine which simultaneously removes and returns blood with anticoagulant citrate dextrose solution A (ACD-A). We aimed at a fluid balance of zero, and no other substitution fluid except FFP and necessary anticoagulant volume was given. To avoid hypocalcaemic episodes, the treatment was run over an approximately 2-hour period allowing for two treatments (of different patients) per day per machine. The flow was between 20 and 60 ml/min, targeting 40-50 ml/min.

### Blood plasma exchange procedures, maintenance

The first five patients included in the treatment protocol were offered maintenance treatment, which was given using the same procedure with three bags of FFP with the Spectra Optia machine at irregular intervals of every 1-6 months for up to one year.

### Blood plasma exchange procedures, lower intensity, longer duration

The long duration low dose procedure was conducted using the Aurora® plasmapheresis machines (using kits Plasmacell 6R2278, Fresenius Kabi), with sodium citrate 4% (ratio 6) as anticoagulant with simultaneous intravenous FFP in the other arm. One bag (610 ml) of FFP was substituted twice per week for 18 weeks. In a few cases, if a treatment session was difficult to arrange, the previous or next treatment was substituted with one plasma exchange with the Spectra Optia machine with two to three bags FFP (each 610 ml) to accommodate for the gap between treatments and achieve a similar average dose of plasma exchange over time.

### Logistics

Patients received an approximate plan for their treatment course (except for maintenance treatments) at the beginning of the treatment protocol. These plans were regularly adjusted, and the patients usually received a final confirmation for the next treatment 1-2 weeks in advance. Reminders were sent to some patients and their relatives on text messages the day before treatment.

### Magnetic resonance imaging (MRI) acquisition and analysis

Participants were scanned at Oslo University Hospital on a 3T GE SIGNA Premier MRI scanner using a 48-channel head coil. 3D T1-weighted anatomical data were collected using an MPRAGE sequence with parameters: repetition time (TR) = 2.526 s, echo time (TE) = 2.836ms, field of view (FOV) = 256 mm, flip angle = 8°, slice thickness = 1 mm, locations per slab = 196 (no overlaps). The T1w images were processed using Freesurfer v.7.4.1 [14], using the longitudinal pipeline, and left and right hemisphere hippocampal volumes were then extracted for further statistical analysis. Cerebral blood flow was assessed using 3D Arterial Spin Labeling (eASL) with spiral acquisition (FOV = 24.0 cm, 4.0 mm slice thickness).

The MRI protocol also included a 3D T2 Cube sequence (TR = 3200 ms, TE = 80 ms, FOV = 25.6 cm, 1.0 mm slice thickness), a 3D Cube T2 FLAIR (TR = 7000 ms, TE = 110 ms, TI = 1978 ms, FOV = 24.0 cm), diffusion-weighted data collected using a 2D Spin Echo Echo-Planar Imaging (EPI) with both AP (TE = 83.5 ms) and PA (TE = minimum) phase-encoding polarities to facilitate distortion correction (TR=4750 ms, FOV=24.0 cm, 1.7 mm slice thickness, and a HyperBand slice acceleration factor of 3), resting-State fMRI acquired with a 2D Gradient Echo EPI sequence (TR = 800 ms, TE = 30 ms, Flip Angle = 52°, FOV = 21.6 cm) and a HyperBand factor of 6, and Quantitative Susceptibility Mapping (QSM) performed using a 3D multi-echo Spoiled Gradient Recalled (SPGR) sequence (10 echoes, TE = Min Full, Flip Angle = 15°, FOV = 25.6 cm) with Asset phase acceleration (factor = 2.0).

### Patient testing during and after treatments. Biobanking

Patients underwent a comprehensive cognitive test battery by trained personnel before, during, and after the intervention. In this manuscript we show data from patient assessment tools Montreal Cognitive Assessment (MoCA) [15], The Informant Questionnaire on Cognitive Decline in the Elderly (IQCODE) (completed by the patient’s next of kin) [16] and a Perceived Research Burden Assessment (PeRBA) [17] test to assess the perceived burden of participation in the treatment protocol. We used Norwegian versions of these tests (Supplementary X). MoCA was administered by certified examiners. Patients were also assessed using the Trail Making Test parts A and B, EQ-5D quality of life questionnaire and delayed recall test from the Consortium to Establish a Registry for Alzheimer’s Disease (CERAD) Word List Memory (10-item word test), but these data are not presented here.

We performed a selected set of physiological measurements aimed at measuring aging processes in patients. As a guide, we were inspired by published testing regimes related to quantifying frailty, and we selected a set of measurements that we could reliably capture in our clinical setting. These measurements were collected at a minimum before and after the main course of treatment, but we aimed to collect them several times before the start of treatment and at several follow-up visits. In this manuscript, we present data on grip strength [18] and forced expiratory volume (FEV1) [19]. We also collected information on blood pressure, pulse rate, oxygen saturation, body temperature, balance standing up by several methods, chair sitting and standing up (5 times) speed, and reaction time to capture a falling ruler, but these measurements are not presented in this manuscript. Additionally, some patients were assessed by cardiopulmonary exercise testing (VO_2_max) protocol before and after treatment and some patients wore a wrist activity tracker for a period of time, but these two metrics were challenging to consistently capture in this study population and the data are incomplete.

Biobanking was performed at a minimum before the first treatment and after the last treatment. Follow-up visits have also included biobanking when possible. Several patients have several baseline biobanked samples from before the treatment started and samples taken during the treatment cycles. EDTA whole blood, EDTA plasma and serum have been collected in all biobank visits while PBMC and citrate plasma have been collected at some visits.

## Results

### Development of plasma exchange protocols for interstitial fluid rejuvenation

A key aim of this project was to develop a plasma exchange protocol that maximizes rejuvenation of the interstitial fluid (Figure 1a). The interstitial fluid (∼12 L) is in equilibrium with blood plasma (∼3 L), with transcapillary exchange rates for albumin and IgG of 5.2%/h and 3%/h, respectively [20–22]. Based on these rates, we estimated that most proteins equilibrate within 48 hours, informing our intensive protocol of ∼2 L plasma exchange every 2-3 days (Figure 1b). To evaluate whether a lower-resource approach could achieve comparable exposure, we also created a less intensive modality exchanging similar total volumes over a longer period.

**Figure 1:**
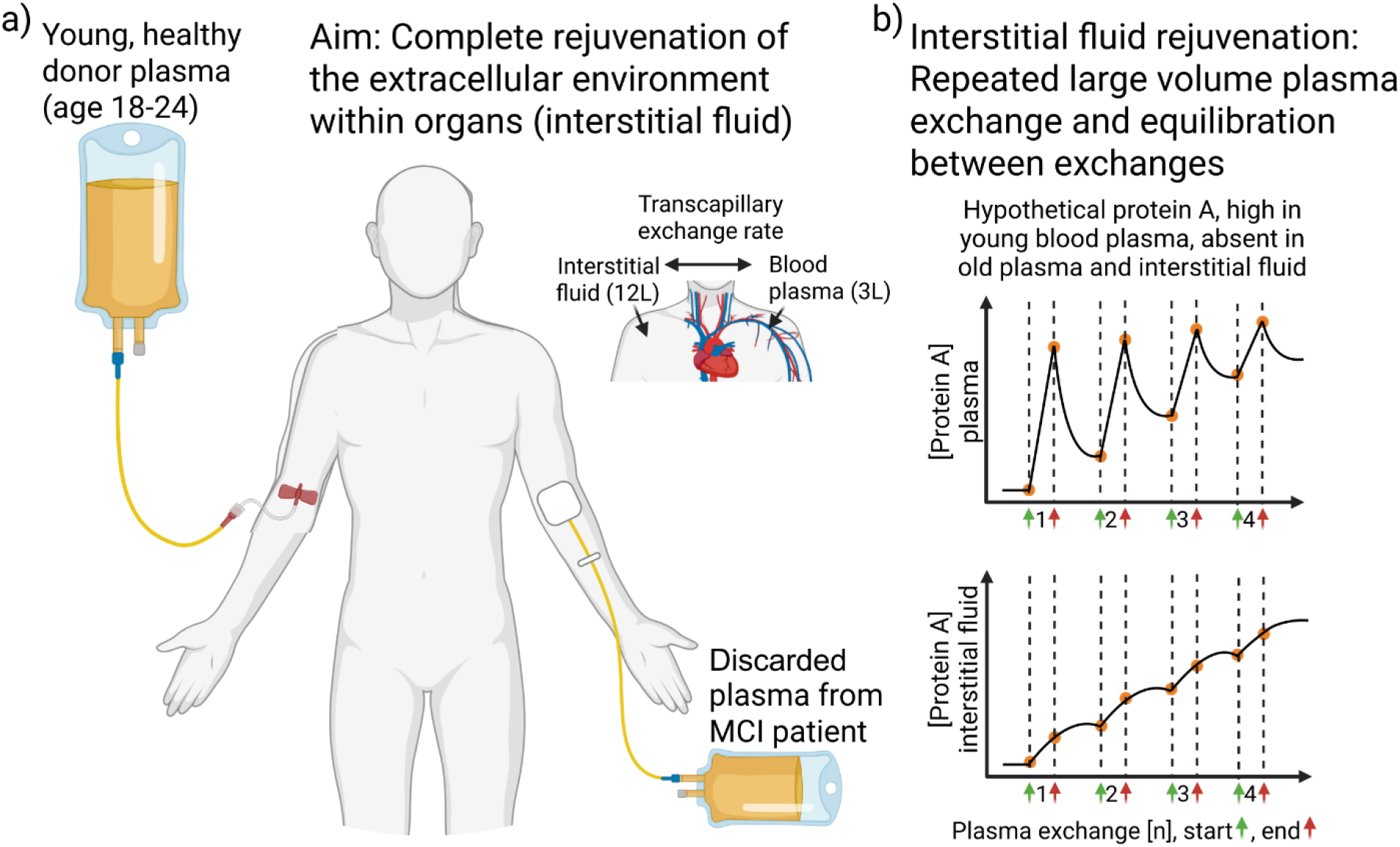
Graphical abstract of our pilot study. The goal is to rejuvenate the interstitial fluid (a), through regularly repeated high-dose plasma exchange (b).

For this pilot study, twelve MCI patients were recruited (Table 1). The first five patients were part of a mixed treatment group where several treatment modalities were explored and protocols refined (Figure 2). The first four patients were started with very low doses of plasma exchange while keeping considerable time between treatments to thoroughly monitor safety in a “pilot-pilot” phase. We then applied an intensive phase of treatment where the goal was to achieve rejuvenation of the interstitial fluid. These patients in the mixed group were also given varying amounts of maintenance treatment after the intensive treatment.

**Table 1:** Baseline characteristics of study participants.

|  | Mixed<br>(n=5) | Intensive<br>(n=4) | Less intensive<br>(n=3) | Total<br>(N=12) |
| --- | --- | --- | --- | --- |
| N | 5 | 4 | 3 | 12 |
| Age, years | 65.4 [64.0–73.8] | 66.9 [61.6–72.5] | 61.8 [57.6–64.1] | 64.7 [61.6–72.5] |
| Sex, male | 3 (60%) | 3 (75%) | 2 (67%) | 8 (67%) |
| MoCA (0–30) | 21.0 [20.0–24.0] | 23.5 [22.8–24.2] | 21.0 [19.0–24.5] | 22.5 [20.8–24.0] |
| IQCODE sum | 57.0 [55.0–70.0] | 55.5 [54.0–57.5] | 54.0 [51.5–55.0] | 55.5 [54.0–57.5] |

Values are median [IQR] or n (%). MoCA is Montreal Cognitive Assessment, IQCODE is Informant Questionnaire on Cognitive Decline in the Elderly

**Figure 2:**
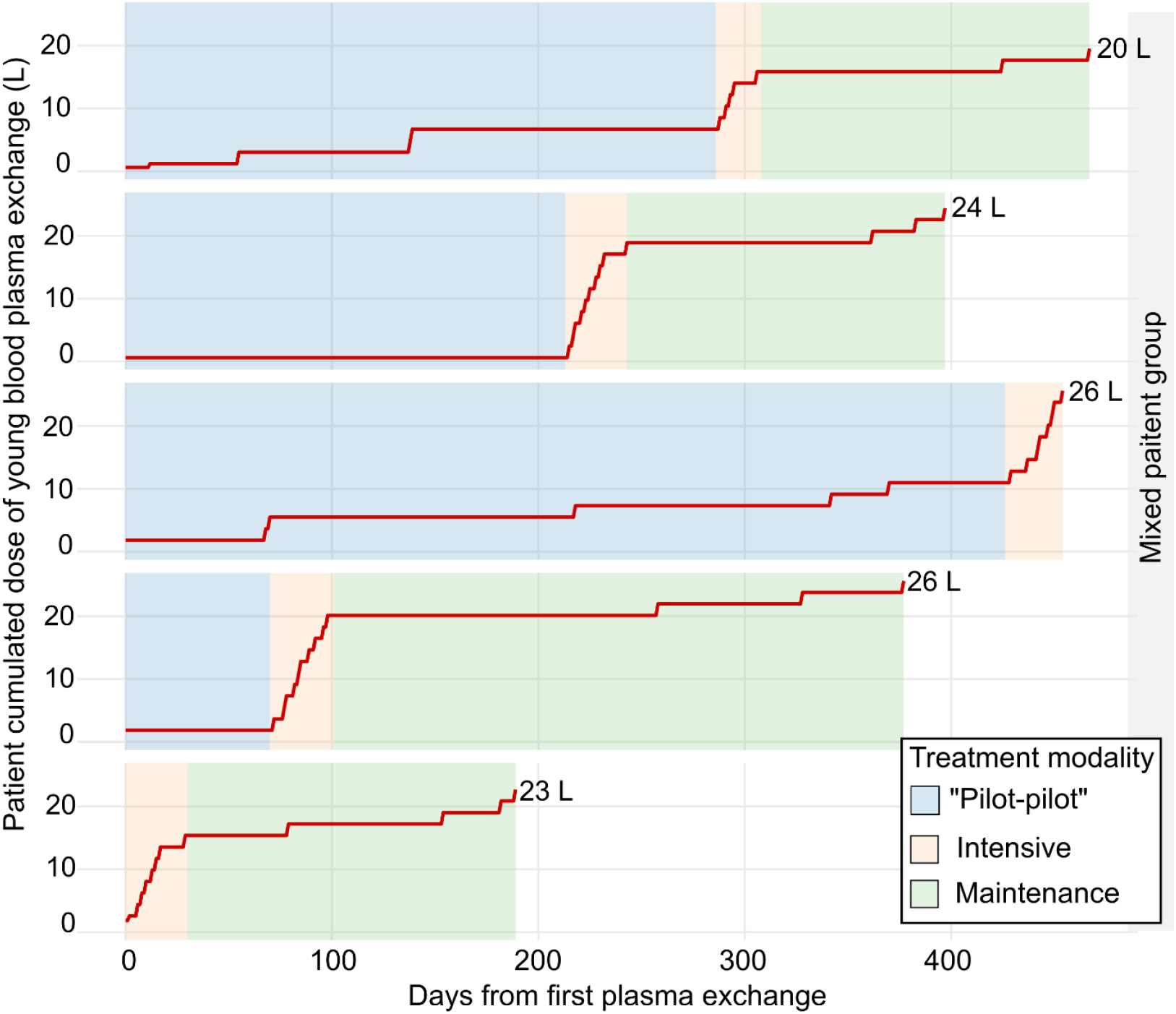
The first five patients in our study were in a “mixed” group, receiving varying doses of the treatment over varying time-intervals. Each red line is for one patient and the number to the right shows the final cumulated plasma exchange for the patient.

After establishing a reliable protocol for how to execute the intensive treatment, four patients were given only the intensive treatment for an average duration of 23.8 days, exchanging on average 17.4 L of blood plasma during this period, an average of 0.5 L per day (Figure 3, Table 2). Three other patients were given a less intensive treatment modality which had an average duration of 83.7 days, over which 16.1 L of blood plasma was exchanged, giving an average of 0.12 L exchange per day (Figure 3, Table 2).

**Table 2:**
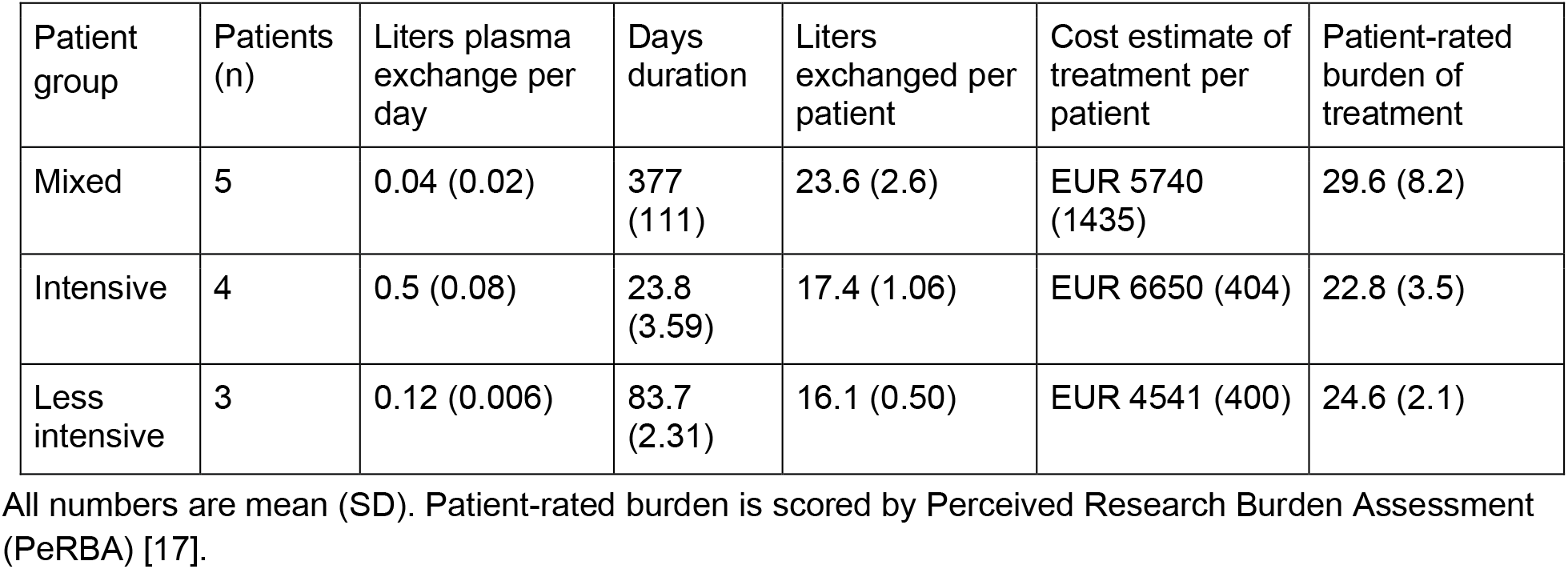
Comparing the three patient groups on metrics of treatment dose, length of treatment, cost of treatment and patient-rated burden of treatment.

**Figure 3:**
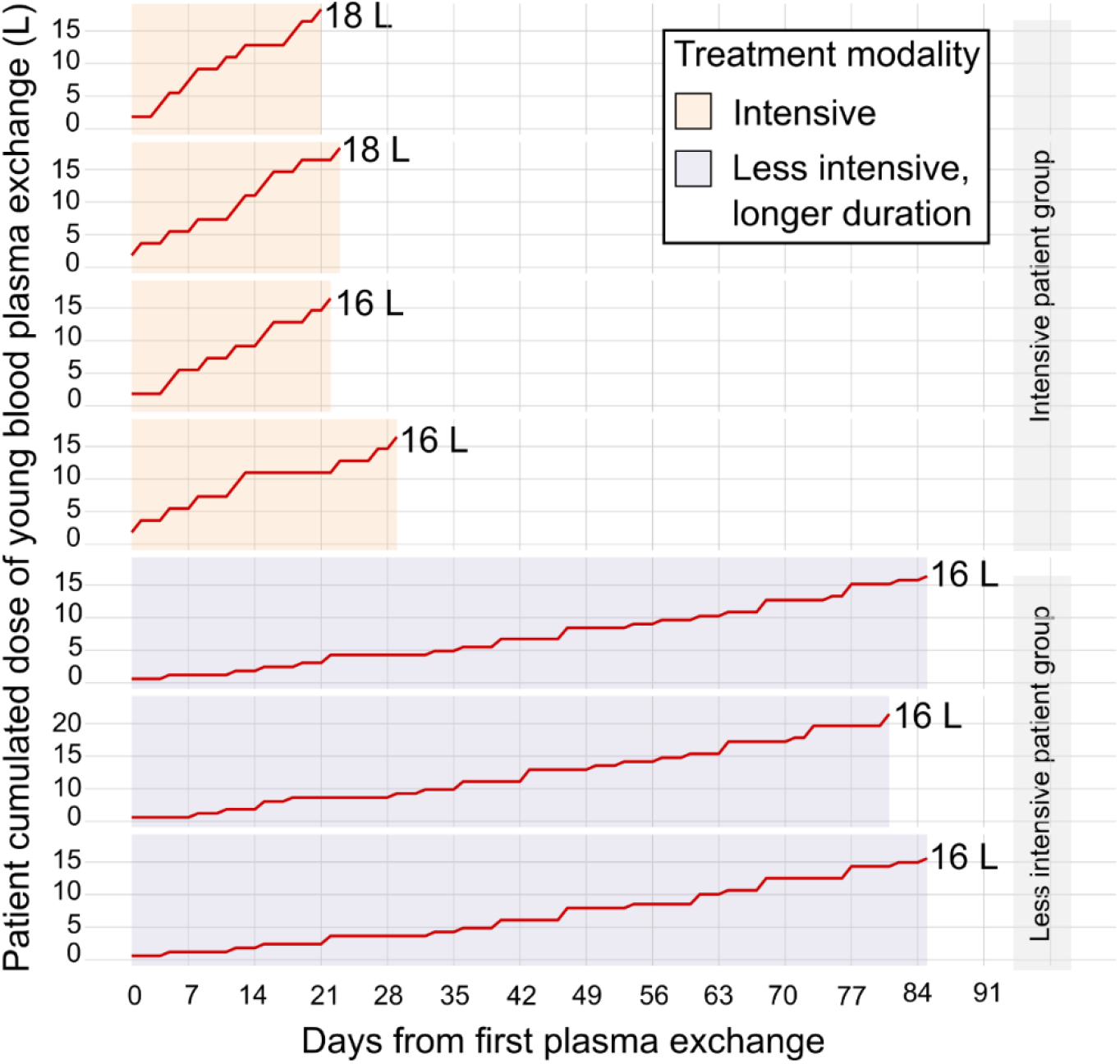
Cumulated dose of plasma exchange for patients in the intensive and less intensive patient groups. Each red line is for one patient and the number to the right of the line shows the final cumulated plasma exchange for the patient.

These treatments involve many visits to the study clinic and many hours spent in a chair during plasma exchange. We monitored the perceived burden of treatment on the patient using a patient burden questionnaire that rates the relative burden on a scale of 21 to 105 points [17]. The treatment modality that was most favorably rated in this context was the intensive treatment (Table 2), where most patients reported that the treatment itself was a very mild burden, despite considerable patient commitment. The mixed patient group and less intensive patient group also reported favorable burden scores (Table 2), but higher than for the intensive treatment. We also created approximate estimates of the cost of the treatment for each patient where we include a price for the donor plasma, consumables, and personnel. The intensive treatment modality is relatively more costly than the others, mostly driven by more personnel costs for intensive procedures (Table 2).

### Adverse events associated with young-donor plasma exchange

Two grade four (life threatening or associated with increased morbidity) adverse events were registered (Table 3). One patient was admitted to hospital 7 days after nine treatments in the intensive phase with a severe infection and probable liver abscess as well as a fast supraventricular tachycardia that the patient had also experienced before treatment. The patient received long-term antibiotic treatment and started heart failure medication. This episode was judged as having a low probability of being caused by the plasma exchange treatment.

**Table 3:**
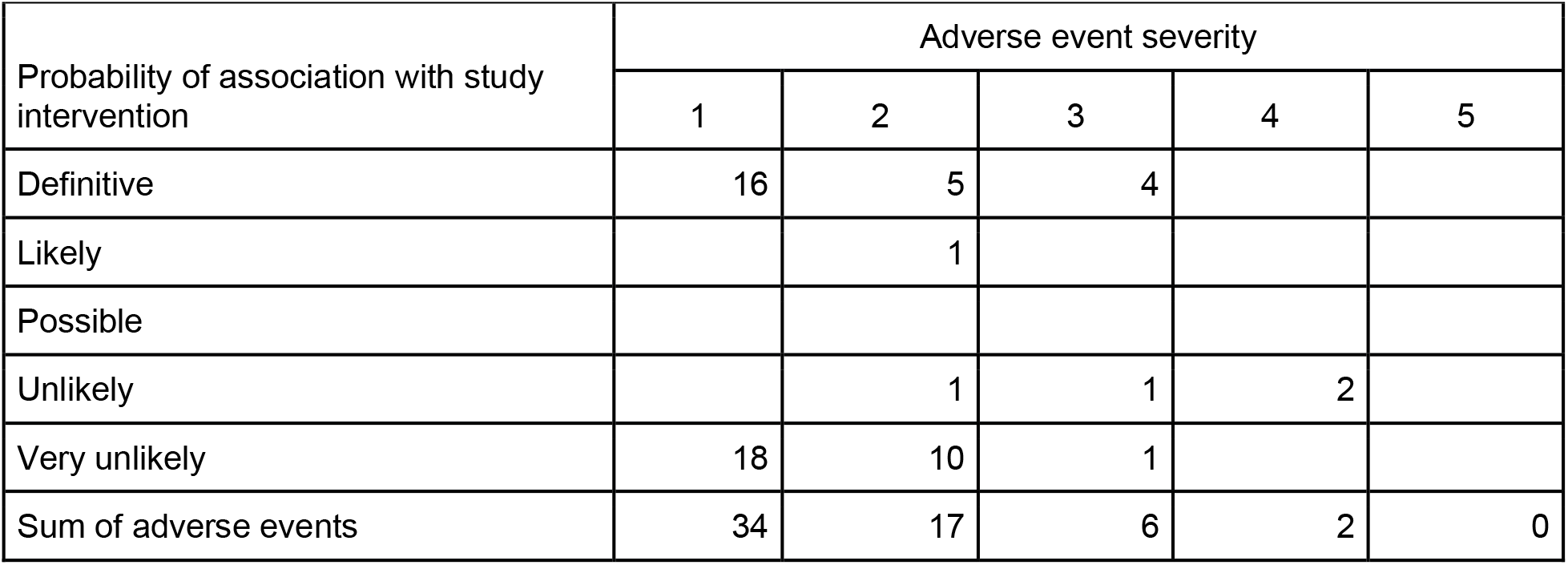
Adverse events and their probability of association with the intervention.

The other grade four adverse event was a patient that noticed a tumor in the arm during treatment that was diagnosed as a rare malignant connective tissue tumor (undifferentiated pleomorphic sarcoma) only weeks after treatment. The tumor was treated with radical excision and radiation, and there has been no evidence of recurrence at the time of writing, 12 months after the operation. The treating oncologist does not believe that the tumor was caused by the treatment, but the possibility cannot be excluded and was judged to have a low probability of being caused by the plasma exchange treatment.

Six grade three adverse events were registered among six different patients (Table 3). Three were generalized urticaria that resulted in abortion of the ongoing treatment and were treated with antihistamine (one tablet) and oral corticosteroids (oral prednisolone 5-10 mg x 1-2). Patients with grade three allergic reactions were observed for 1-2 hours until their condition started to improve and were sent home to be observed by their relatives. They all received prednisolone tablets to use at home and had fully recovered the next day. One case of cough without dyspnea resulted in discontinuation of the ongoing bag of plasma, but continuation of the treatment the same day and treatment with antihistamine and oral prednisolone. Another grade three adverse event was a patient diagnosed with a paroxysmal atrial flutter during a break in the procedure after five treatments. The patient was given anticoagulant treatment, and the episode was deemed unlikely to be associated with the treatment. Finally, one patient had a grade three adverse event caused by one of the physical tests in the study.

Seventeen grade two adverse events were registered among eight different patients, of which six had a probable or likely association with the treatment. These included vivid dreams, mild/localized urticaria (two cases), cough, and two cases of mild symptomatic hypocalcemia. In addition, one patient had a mildly elevated Troponin T and creatine kinase after a VO_2_max test. In one patient, a mild anemia that had lasted for several months before treatment was acknowledged after five treatments. In this case, treatments were postponed until diagnostic workup was completed, and the patient was cleared to continue by the gastroenterologist (during this period, an atrial flutter was also found, see above). Two other patients also had supraventricular arrhythmia, but both had also experienced this before treatment.

Finally, 34 grade one adverse events in twelve different patients (all patients had at least one reported adverse event). Of these, 16 had a probable or likely association with the study intervention (among nine different patients). These included likely supraventricular extrasystoles, increased sleepiness in the evening, mild cough/dry throat, feeling of cold and mild (0.6 degrees) increased temperature during treatment, feeling unwell the evening before treatment, feeling warm before bedtime, itchy eyes during treatment (three times, same patient), increased number of skin spots, itchy face during treatment, itchy skin without urticaria, dizziness after the procedure, three cases of mild hypocalcemia. One significant hematoma due to venous access was noted.

### Preliminary measurements of cognitive performance, physiological aging and brain metrics before and after young-donor plasma exchange

Most patients we recruited were included with MoCA scores in the range of 20-25, typical of an MCI population. Two patients started with MoCA scores of 17, and both demonstrated further cognitive decline, as reflected by drop in MoCA scores and a concomitant decline in functional status after treatment in our study (Figure 4). We also collected the Informant Questionnaire on Cognitive Decline in the Elderly (IQCODE) measurements, a questionnaire given to informants (the spouse or a relative) that aims to capture changes in the capacity of the patient to handle sixteen everyday life tasks compared to ten years previously (Figure 4).

**Figure 4:**
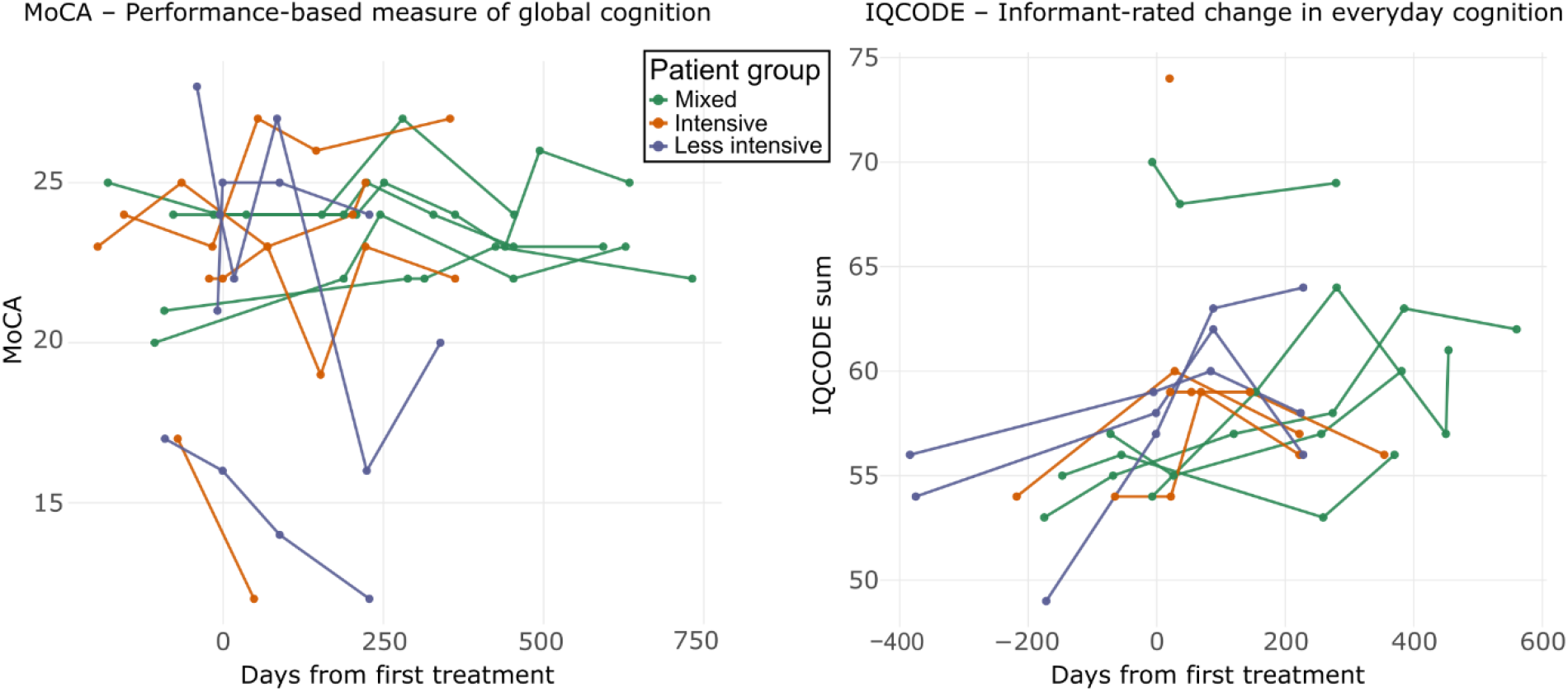
Cognitive test results for all pilot study patients. Each patient has one line, colored by the treatment group they were in. MoCA quantifies patient cognitive performance (higher is better). IQCODE quantifies the patient’s closest relative’s judgement of the patient’s cognitive decline related to everyday function. IQCODE scores are between 16-80 where higher is worse and 48 means no overall change from ten years earlier.

Because our treatment protocol is targeting aging processes, we were eager to follow aging-related metrics in patients that are not necessarily related to MCI or dementia. We composed a set of physiological tests, inspired by frailty testing, but modified to fit our clinical environment.

## Discussion

In this pilot study, we treated twelve patients with MCI due to AD using large-volume young-donor plasma exchange across three different protocols. All patients completed their assigned treatment, and we did not observe safety signals warranting concern.

### The strengths of our young-donor plasma exchange protocol

Previous human studies have either removed patient plasma or added young plasma, but studies combining the two have not been published. The AMBAR trial exchanged 3–4.5 L of patient plasma per session and replaced it with a saline and albumin solution [7]. The PLASMA study infused ∼250 mL of young donor plasma weekly for four weeks without removing old plasma [9], and the GRF6019 trial infused 100–250 mL daily for five days of a proprietary young plasma fraction [8]. The ongoing Norwegian ExPlas trial uses 200 mL infusions of plasma from exercise-trained young donors in patients with MCI or early AD, also without plasma removal [23]. Our protocol is the first to combine large-volume removal of patient plasma with replacement by young donor plasma, addressing both sides of the heterochronic blood exchange equation. This is the first published protocol that can realistically change the signaling environment within organs to a younger state.

### Safety and feasibility

While two patients had severe adverse events, the probability of them being related to the procedure was considered small. Adverse events attributed to the study protocol were limited to known adverse events of plasma exchange, including generalized urticaria which affected three of twelve patients in this pilot study. Generally, our rates of adverse events were similar to what was seen in the larger AMBAR study [7]. Our safety findings support a larger clinical trial based on our pilot study protocol.

Patients with MCI and probable AD have no effective treatments available, poor long-term prognosis and a reduced cognitive ability to judge the risks of participating in research projects. This dilemma was discussed in our group’s patient board and with the Ethics Committee during the design of the study and was addressed by involving the nearest relative in the consent process. We believe that this contributed to a highly motivated and informed group of patients and relatives in the study and no dropouts. The reduced cognitive ability of the patients did not cause significant problems for the logistics of the study visits.

In this study, we present a protocol designed to get as close as practically possible to interstitial fluid rejuvenation through plasma exchange in humans. When the goal is to rejuvenate the interstitial fluid, large volumes of young donor blood plasma will be required. Generally, 3+12 L (blood plasma + interstitial fluid volume [22]) should be exchanged to achieve an environment within organs that can be assumed to be rejuvenated. The most obvious challenge with this protocol is the access to young donor blood plasma. Donors in this age group have high attrition rates and are often deferred due to lifestyle factors such as travel and new sexual partners.

Plasma donation is also relatively time consuming, limited by access to plasmapheresis machines and qualified personnel. Here, we have relied on a single blood center for access to blood plasma, but a larger trial requiring an increased number of plasma units would require several blood centers.

### Implications for patient selection in future trials

Two patients in our cohort entered the study with MoCA scores of 17, just above the exclusion threshold. Both experienced further cognitive decline during the follow-up period (Figure 4), accompanied by loss of functional status. We cannot distinguish whether this decline reflects natural disease progression or a lack of treatment benefit in patients with more advanced disease. Either way, the observation suggests that targeting aging processes may be less helpful once substantial brain damage has accumulated. Future plasma exchange studies in MCI patients may benefit from a more stringent cognitive threshold at enrollment, for example MoCA ≥ 20.

### Exploratory outcomes on aging biology and brain structure

Beyond cognitive outcomes, we collected physical and brain measurements that change with aging (Figure 5). Grip strength predicts mortality and disability in older adults [18], and FEV1 is a marker of biological aging pace [19]. Importantly, an analogue measurement of grip strength in rodents is improved by heterochronic parabiosis [1]. Hippocampal volume is reduced in AD progression [24], and cerebral blood flow declines with both aging and AD [25]. We were able to collect all four metrics across the treatment course, supporting their inclusion as secondary outcomes in a future controlled trial.

**Figure 5:**
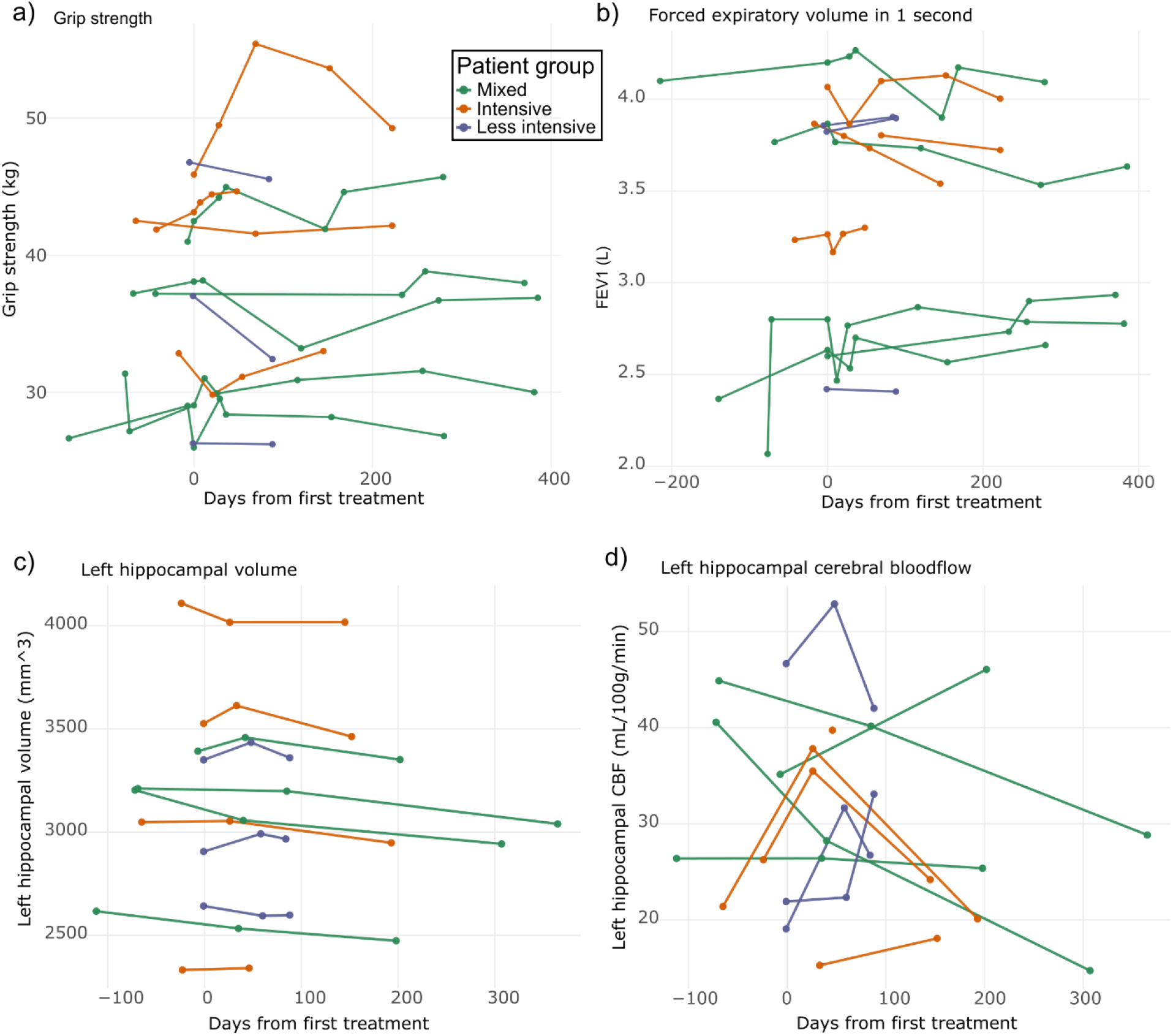
Tests of patient physical function and brain physiology. Each patient has one line, colored by the treatment group they were in. a,b) For grip strength, we computed the mean of three measurements of each hand. For FEV, we used the mean of three measurements. c,d) Magnetic resonance measurements of brain left hippocampal volume and cerebral blood flow.

### Study limitations

Our pilot study has a relatively small sample size of N=12 in an open, unblinded study design. Heterogenous protocols across three treatment groups further limit the N in each unique treatment protocol. The relatively extensive treatment and medical care patients receive in this pilot study creates potential for placebo effects. The study design does not allow inference related to treatment efficacy. A longer-term follow-up including a higher number of patients is required to determine whether the cognitive trajectories observed reflect a true treatment effect, given the typically slow and variable course of AD progression.

### Concluding remarks: Is cognitive decline the best target when treating aging?

While aging is an obvious contributor to a large group of diseases, designing clinical trials to delay or prevent disease is complicated by the complexities of exactly how aging contributes. Here, we focused on MCI with the hypothesis that aging is a contributor to the processes that drive cognitive decline in MCI patients. While there are strong links between aging and cognitive performance and demonstrated improvement in cognitive performance in older rodents exposed to heterochronic parabiosis, it may be that the disease progression of MCI is not affected by modulation of aging processes. The interstitial fluid rejuvenation protocol established here should be relevant to modulate aging processes in any organ. We speculate that frailty is an especially good target for this treatment protocol based on rodent data demonstrating simultaneous muscle and cognitive improvement [10]. A future application of our study protocol could leverage this by targeting more frail MCI patients, potentially improving both the frail state and the brain and benefitting from synergy due to the strong relationship between frailty and dementia [26]. Our study protocol should be freely adaptable to any age-related disease. We conclude that the safety profile and feasibility of this treatment protocol demonstrates that it is acceptable for testing in a larger, randomized study of treatment efficacy.

## Acknowledgments

The authors thank patients, their families and the young plasma donors for their generous and sustained contribution to the study.

## Declarations

### Ethics approval and consent

The study was approved by the Regional Committee for Medical and Health Research Ethics, South-East Norway, Subcommittee A application reference number 520697. It was conducted in accordance with the Declaration of Helsinki. All data were processed according to EU/GDPR regulations. All patients provided written informed consent (Supplementary X) prior to enrollment, co-signed by their closest relative. All blood plasma donors provided written informed consent (Supplementary X) prior to blood plasma donation.

### Trial registration

ClinicalTrials.gov NCT06234436 (registered 22 December 2023).

### Data availability

The datasets generated and analyzed during this study contain personal health information from a small clinical cohort and cannot be made publicly available due to patient privacy considerations and Norwegian/EU data protection regulations (GDPR). De-identified data may be made available to qualified researchers upon reasonable request to the corresponding author and subject to approval from the Norwegian Regional Ethics Committee and a data transfer agreement.

### Author contributions

Patient treatment and sample collection was performed by Arne Søraas, Lana Nagmadin Karim, Elena Danilova, Mette Stausland Istre, Sara Nygaard, Tone Reinton Utgård, Sandra Ceprnjic, Olaug Reiakvam, Elisabeth Edvardsen, Lars T. Westlye, Lise Sofie Haug Nissen-Meyer, Håkon Ihle-Hansen and Petter Holland. Conceptual planning, material preparation, data collection and analysis were performed by Arne Søraas, Andreas Engvig, Dag Alnæs, Anders B. Nygaard, John Arne Dahl, Lise Sofie Haug Nissen-Meyer and Petter Holland. The first draft of the manuscript was written by Arne Søraas and Petter Holland and all authors commented on subsequent versions of the manuscript. All authors read and approved of the final manuscript.

## References

1. Conboy IM, Conboy MJ, Wagers AJ, et al (2005) Rejuvenation of aged progenitor cells by exposure to a young systemic environment. Nature 433:760–764. 10.1038/nature03260

2. Villeda SA, Luo J, Mosher KI, et al (2011) The ageing systemic milieu negatively regulates neurogenesis and cognitive function. Nature 477:90–96. 10.1038/nature10357

3. Villeda SA, Plambeck KE, Middeldorp J, et al (2014) Young blood reverses age-related impairments in cognitive function and synaptic plasticity in mice. Nature Medicine 20:659–663. 10.1038/nm.3569

4. Castellano JM, Mosher KI, Abbey RJ, et al (2017) Human umbilical cord plasma proteins revitalize hippocampal function in aged mice. Nature 544:488–492. 10.1038/nature22067

5. Smith LK, He Y, Park J-S, et al (2015) β2-microglobulin is a systemic pro-aging factor that impairs cognitive function and neurogenesis. Nat Med 21:932–937. 10.1038/nm.3898

6. Holland P, Istre M, Ali MM, et al (2024) Epigenetic aging of human blood cells is influenced by the age of the host body. Aging Cell 23:e14112. 10.1111/acel.14112

7. Boada M, López OL, Olazarán J, et al (2020) A randomized, controlled clinical trial of plasma exchange with albumin replacement for Alzheimer’s disease: Primary results of the AMBAR Study. Alzheimer’s and Dementia 16:1412–1425. 10.1002/alz.12137

8. Hannestad J, Koborsi K, Klutzaritz V, et al (2020) Safety and tolerability of GRF6019 in mild-to-moderate Alzheimer’s disease dementia. Alzheimer’s & Dementia: Translational Research & Clinical Interventions 6:e12115. 10.1002/TRC2.12115

9. Sha SJ, Deutsch GK, Tian L, et al (2019) Safety, Tolerability, and Feasibility of Young Plasma Infusion in the Plasma for Alzheimer Symptom Amelioration Study: A Randomized Clinical Trial. JAMA Neurol 76:35–40. 10.1001/jamaneurol.2018.3288

10. Rebo J, Mehdipour M, Gathwala R, et al (2016) A single heterochronic blood exchange reveals rapid inhibition of multiple tissues by old blood. Nature Communications 7:1–11. 10.1038/ncomms13363

11. Winblad B, Palmer K, Kivipelto M, et al (2004) Mild cognitive impairment – beyond controversies, towards a consensus: report of the International Working Group on Mild Cognitive Impairment. Journal of Internal Medicine 256:240–246. 10.1111/j.1365-2796.2004.01380.x

12. Nissen-Meyer LSH, Hervig T, Fevang B, et al (2022) COVID-19 convalescent plasma from Norwegian blood donors. Tidsskrift for Den norske legeforening. 10.4045/tidsskr.22.0057

13. Steinsvåg CT, Espinosa A, Flesland Ø (2013) Eight years with haemovigilance in Norway. What have we learnt? Transfusion and Apheresis Science 49:548–552. 10.1016/j.transci.2013.09.013

14. Fischl B (2012) FreeSurfer. NeuroImage 62:774–781. 10.1016/j.neuroimage.2012.01.021

15. Nasreddine ZS, Phillips NA, Bédirian V, et al (2005) The Montreal Cognitive Assessment, MoCA: A Brief Screening Tool For Mild Cognitive Impairment. Journal of the American Geriatrics Society 53:695–699. 10.1111/j.1532-5415.2005.53221.x

16. Jorm AF, Jacomb PA (1989) The Informant Questionnaire on Cognitive Decline in the Elderly (IQCODE): socio-demographic correlates, reliability, validity and some norms. Psychol Med 19:1015–1022. 10.1017/s0033291700005742

17. Lingler JH, Schmidt KL, Gentry AL, et al (2014) A New Measure of Research Participant Burden: Brief Report. Journal of Empirical Research on Human Research Ethics 9:46–49. 10.1177/1556264614545037

18. Bohannon RW (2019) Grip Strength: An Indispensable Biomarker For Older Adults. CIA 14:1681– 1691. 10.2147/CIA.S194543

19. Belsky DW, Caspi A, Houts R, et al (2015) Quantification of biological aging in young adults. Proceedings of the National Academy of Sciences 112:E4104–E4110. 10.1073/pnas.1506264112

20. Parving H-H, Rossing N (1973) Simultaneous Determination of the Transcapillary Escape Rate of Albumin and IgG in Normal and Long-Term Juvenile Diabetic Subjects. Scandinavian Journal of Clinical and Laboratory Investigation 32:239–244. 10.3109/00365517309082466

21. Parving HH, Jensen HÆ, Westrup M (1977) Increased transcapillary escape rate of albumin and IgG in essential hypertension. Scandinavian Journal of Clinical and Laboratory Investigation 37:223–227. 10.3109/00365517709091486

22. Hall J, Hall M (2020) Guyton and Hall Textbook of Medical Physiology, 14th ed. Elsevier, Philadelphia, PA

23. Tari AR, Berg HH, Videm V, et al (2022) Safety and efficacy of plasma transfusion from exercise-trained donors in patients with early Alzheimer’s disease: protocol for the ExPlas study. BMJ Open 12:e056964. 10.1136/bmjopen-2021-056964

24. Jack CR, Petersen RC, Xu YC, et al (1999) Prediction of AD with MRI-based hippocampal volume in mild cognitive impairment. Neurology 52:1397–1403. 10.1212/wnl.52.7.1397

25. Graff BJ, Harrison SL, Payne SJ, El-Bouri WK (2023) Regional Cerebral Blood Flow Changes in Healthy Ageing and Alzheimer’s Disease: A Narrative Review. Cerebrovasc Dis 52:11–20. 10.1159/000524797

26. Engvig A, Maglanoc LA, Doan NT, et al (2023) Data-driven health deficit assessment improves a frailty index’s prediction of current cognitive status and future conversion to dementia: results from ADNI. GeroScience 45:591–611. 10.1007/s11357-022-00669-2

